# The Association between Early Country-level Testing Capacity and Later COVID-19 Mortality Outcomes

**DOI:** 10.1101/2021.01.18.21249998

**Authors:** Sneha Kannoth, Sasikiran Kandula, Jeffrey Shaman

## Abstract

**Background:** The COVID-19 pandemic has overrun hospital systems while exacerbating economic hardship and food insecurity on a global scale. In an effort to understand how early action to find and control the virus is associated with cumulative outcomes, we explored how country-level testing capacity affects later COVID-19 mortality.

**Methods:** We used the Our World in Data database to explore testing and mortality records in 27 countries from December 31, 2019 to September 30, 2020; we applied ordinary-least squares regression with clustering on country to determine the association between early COVID-19 testing capacity (cumulative tests per case) and later COVID-19 mortality (time to specified mortality thresholds), adjusting for country-level confounders, including median age, GDP, hospital bed capacity, population density, and non-pharmaceutical interventions.

**Results:** Higher early testing implementation, as indicated by more cumulative tests per case when mortality was still low, was associated with longer accrual time for higher per capita deaths. For instance, a higher cumulative number of tests administered per case at the time of 6 deaths per million persons was positively predictive of a longer time to reach 15 deaths per million, after adjustment for all confounders (β=0.659; *P*=0.001).

**Conclusions:** Countries that developed stronger COVID-19 testing capacity at early timepoints, as measured by tests administered per case identified, experienced a slower increase of deaths per capita. Thus, this study operationalizes the value of testing and provides empirical evidence that stronger testing capacity at early timepoints is associated with reduced mortality and better pandemic control.

## INTRODUCTION

As of January 18, 2021, the coronavirus disease 2019 (COVID-19) pandemic, caused by infection with severe acute respiratory syndrome coronavirus 2 (SARS-CoV-2), had produced a total of 95.8 million cases, and 2.04 million deaths globally.^1^ COVID-19 has inundated healthcare systems throughout the world with increased demand for clinical resources, including intensive care units, hospital beds, and personal protective equipment (PPE).^2^ The public health crisis has also worsened mental health outcomes by increasing the prevalence of stress, anxiety, and depression worldwide.^3-4^ Furthermore, COVID-19 has and will produce adverse socio-economic outcomes globally as the International Labor Organization projects the pandemic will push 200 million to unemployment, and the World Food Program projects 265 million will face acute hunger because of the pandemic.^5-6^

It is therefore essential to control the spread of SARS-CoV-2. To do so, nations have primarily relied on non-pharmaceutical interventions, including testing, contact tracing, and isolation,^7^ as no universally accepted therapeutic regimen or vaccine was available for this infectious respiratory virus for most of 2020.^8^ Testing, tracing and isolation can be the cornerstones of COVID-19 control policy; however, country-level capacity for recognizing and verifying cases has varied.^9^ Limitations in this capacity may stem from shortages in testing equipment, such as swabs, reagents, and PPE, in addition to having a limited number of qualified personnel. These limitations have the potential to increase case numbers amid an already destabilized testing infrastructure.^10^

There are few studies that have evaluated the effects of testing capacity strength on future COVID-19 outcomes^11-12^. Pan et al. (2020) demonstrated that symptom survey rates in Wuhan, China were indicative of a smaller effective reproductive number of SARS-CoV-2 and were associated with reduced daily confirmed cases.^11^ In contrast, Chaudry et al. (2020) adopted a more global approach and found that wide-spread testing, in addition to full lockdowns, were not associated with country-level COVID-19 death rates.^12^ Given such inconsistencies in the literature, we aimed to explore whether early country-level testing capacity, measured as *tests administered per case identified*, has an association with COVID-19 mortality. The rationale is that a higher level of testing per case identified is indicative of more proactive efforts to find infections in the broader community and break chains of transmission. We assessed COVID-19 mortality as opposed to COVID-19 reported infections, given that testing demand and capacity can change in response to the number of reported infections. In quantifying the effects of testing capacity, researchers may be better able to gauge the value of rapidly scaling up and promoting these public health strategies. Thus, we aimed to assess the relationship between the rigor of early testing and future COVID-19 mortality across countries.

In exploring this research question, we used information regarding daily cumulative testing and mortality for 27 countries.^1^ We also incorporated data regarding the timing of non-pharmaceutical interventions, including confinement, school/work closures, event cancellations, travel restrictions, and health practices (contact tracing and mask wearing).^13^ We hypothesize that higher country-level tests per case in the early stages of the outbreak is associated with a longer time (measured in days) to reach specific mortality thresholds (i.e. Y deaths per million), after adjusting for country-level characteristics, including median age, GDP, hospital bed capacity, population density, and timing of NPI implementation.

## METHODS

### Study design and sample

We used the Our World in Data, a publicly available scientific database, to quantify country-level testing capacity and health outcomes.^1^ The study inclusion criteria broadly identified countries whose first documented testing count was less than 100 tests, and whose case count (at that particular point) was less than the test count. These countries included Bangladesh, Bolivia, Czech Republic, Estonia, Finland, Hungary, Iceland, India, Israel, Italy, Kenya, Latvia, Mexico, Morocco, Nepal, New Zealand, Pakistan, Panama, Paraguay, Poland, Portugal, Serbia, South Africa, South Korea, Switzerland, Tunisia, and the United States (N=27).

### Predictor

#### Early testing capacity

Country-level testing capacity was operationalized as the number of reported tests per confirmed COVID-19 case measured at early mortality thresholds, i.e. X deaths per million, where X ranged from 1 to 25.

### Outcome

#### COVID-19 mortality

COVID-19 mortality was operationalized as the timespan (measured in days) to reach specific country-level mortality thresholds, measured at Y deaths per million, where Y ranged from 2 to 30. The starting point of the timespan was identified as the date corresponding to the early testing capacity measure.

### Confounders

In assessing the association between early testing capacity and mortality outcomes, we accounted for potential country-level confounders, including median age, gross domestic product (GDP), hospital bed capacity (i.e. hospital beds per 1000 people), population density, and non-pharmaceutical intervention (NPI) implementation. NPI categories included 1) mandatory/advised confinement; 2) school/work closures; 3) event cancellations (restrictions on public gatherings; entertainment/cultural sector closures; public services closures); 4) travel restrictions (international travel restrictions; restricted freedom of movement); and 5) health practices (contact tracing; mask wearing). If an NPI category was implemented during the full timespan between a specified date marking early testing capacity and a specified date marking the mortality threshold, then it was assigned a value of 1. The summary NPI variable during this specified time span was expressed as either a sum of the five NPI category values (NPI Sum; range = 0-5) or an average of the five NPI category values (NPI Average; range = 0-1). The summary NPI variable accounted for the time that a NPI was in place, and the number of NPIs that were in place. NPI data was retrieved from IBM Research: Worldwide NPI Tracker for COVID-19.^13^

### Statistical analysis

We determined the trajectory of cumulative tests per case and deaths per million for each country in our analysis from December 31, 2019 to September 30, 2020. We additionally modelled the daily time-evolving pattern of country-level testing capacity and country-level mortality rate, in addition to creating a log-log plot assessing the crude relationship between cumulative tests per case and cumulative deaths per million, measured on September 30, 2020 and determined the corresponding Pearson correlation coefficient. We then used ordinary least-squares (OLS) regression, with clustering on spatial unit (country) to assess the association between early testing capacity and later COVID-19 mortality outcomes. We implemented four models, each adjusting for a different set of covariates. Model 1 adjusted for median age, and gross domestic product (GDP) at the country-level; Model 2 adjusted for median age, GDP, and hospital bed capacity at the country-level; Model 3 adjusted for median age, GDP, hospital bed capacity, and population density at the country-level; Model 4 and 5 adjusted for median age, GDP, hospital bed capacity, population density, and NPIs, either summed or averaged, at the country-level. We used R.version 4.0.3 to perform all analyses.

### Bootstrap analysis

To approximate the variance of the model effect estimates, we conducted 1000 bootstraps on the model coefficients, randomly selecting all 27 countries in the sample, with replacement (Figure Suppl 1A-1E).

## RESULTS

### Global trajectory of testing capacity and COVID-19 mortality rates

Figures 1A and 1B demonstrate the testing capacity and mortality rate trajectory among the 27 specified countries from December 31, 2019, to September 30, 2020. During this time period, we found a crude negative association between log-transformed tests per case and log-transformed deaths per million, r(25) = −0.59, p=0.001 (Figure 2). Time-evolving patterns of country-level testing capacity and country-level mortality indicate that countries with higher testing capacity early in the pandemic experienced a slower accrual of deaths per capita (Figure Suppl 2).

**Figure 1A-B.**
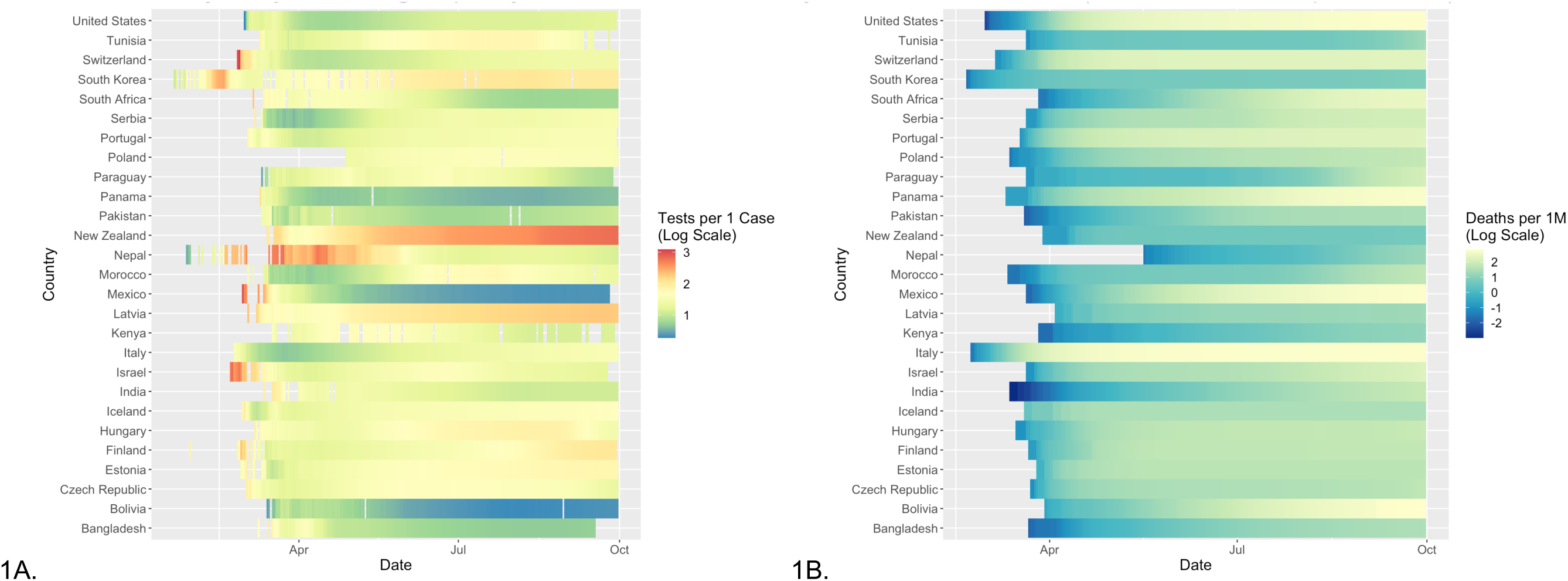
Trajectory of Testing Capacity and COVID-19 Mortality, Across 27 Countries (Dec 31, 2019 – Sept 30, 2020)^a,b^. ^a^Figure 1A. Total Tests per Case for SARS-CoV-2 Over Time (log); ^b^Figure 1B. Total COVID-19 Deaths per Million Over Time (log)

**Figure 2.**
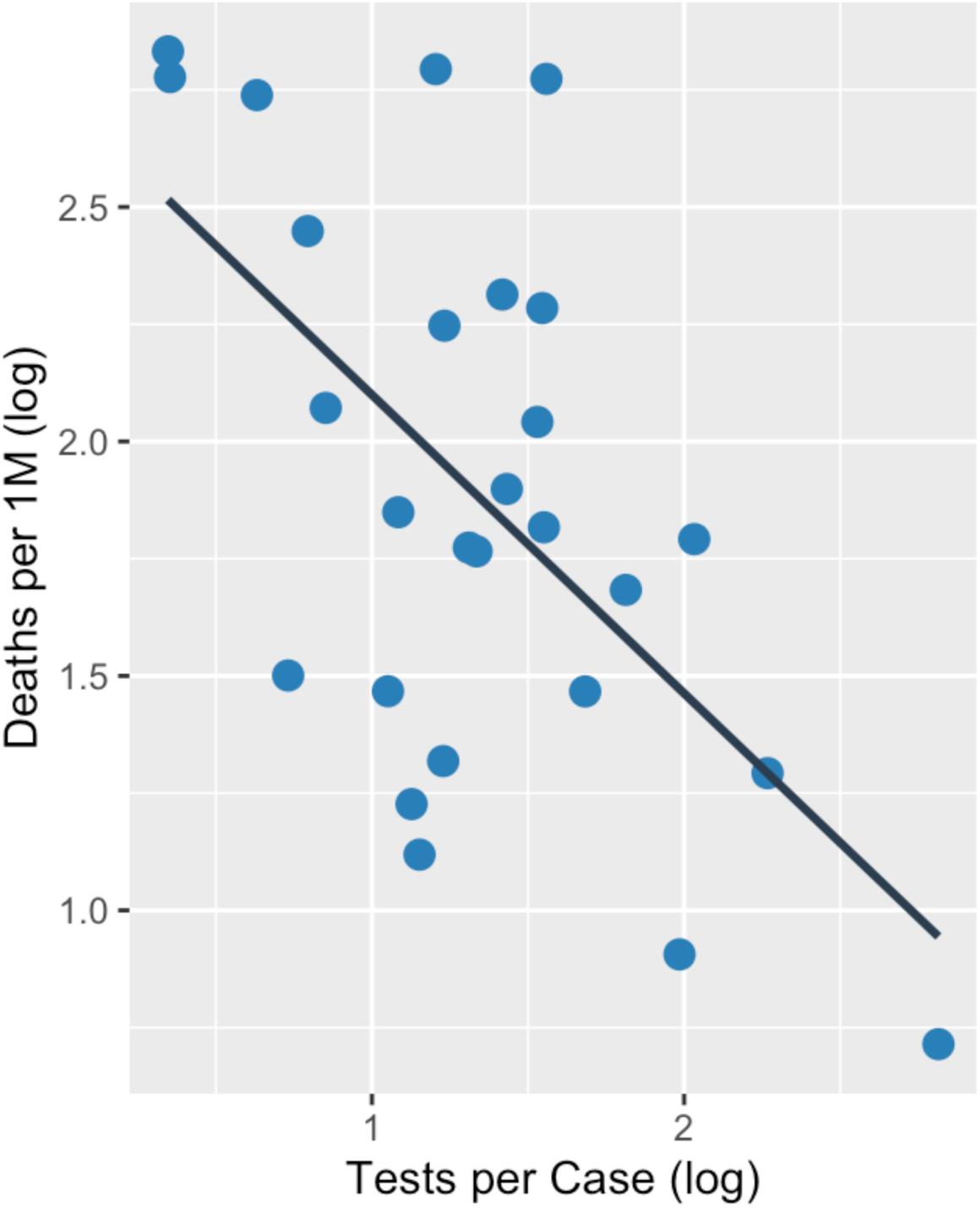
Total COVID-19 Deaths per Million vs. Total Tests per COVID-19 Case on Sept 30, 2020, Across 27 Countries^a,b^. ^a^Each plot point represents an individual country’s total tests per case and total deaths per 1 million ^b^Pearson correlation coefficient: −0.59 (*P* = 0.001)

### Ordinary least-squares regression analyses

Table 1 provides the adjusted effect estimates regarding the association between tests per case at 6 deaths per million and time (days) to 15 deaths per million. We found that the number of tests per case at 6 deaths per million is associated with the time (days) to reach 15 deaths per million, when adjusting for median age and GDP (Model 1: β = 0.544; *P* = 0.004); adjusting for median age, GDP, and hospital bed capacity (Model 2: β = 0.564; *P* = 0.004); adjusting for median age, GDP, hospital bed capacity, and population density (Model 3: β = 0.649; *P* = <0.001); adjusting for median age, GDP, hospital bed capacity, population density, and NPI Sum (Model 4: β = 0.659; *P* = 0.001); and adjusting for median age, GDP, hospital bed capacity, population density, and NPI Average (Model 5: β = 0.659; *P* = 0.001). In all instances, the effect estimates indicate that greater tests per case at 6 deaths per million are positively predictive of a longer time to 15 deaths per million, i.e. a slower accrual of mortality.

**Table 1.** Adjusted Regression Models of Tests per COVID-19 Case at 6 Deaths per 1 Million and Time (Days) to 15 COVID-19 Deaths per 1 Million

|  | *Estimate* | *Standard Error* | *P-value* |
| --- | --- | --- | --- |
| **Model 1** |  |  |  |
| Tests per 1 Case at 6 Deaths per 1 Million | 0.544 | 0.165 | 0.004 |
| Median Age | -0.173 | 0.253 | 0.501 |
| GDP | -0.0003 | 0.0001 | 0.007 |
| **Model 2** |  |  |  |
| Tests per 1 Case at 6 Deaths per 1 Million | 0.564 | 0.169 | 0.004 |
| Median Age | 0.148 | 0.364 | 0.688 |
| Country-Level Gross Domestic Product | -0.0003 | 0.0001 | 0.009 |
| Hospital Beds per 1000 | -1.568 | 1.097 | 0.170 |
| **Model 3** |  |  |  |
| Tests per 1 Case at 6 Deaths per 1 Million | 0.649 | 0.164 | <0.001 |
| Median Age | 0.072 | 0.335 | 0.833 |
| Country-Level Gross Domestic Product | -0.0003 | 0.0001 | 0.021 |
| Hospital Beds per 1000 | -1.127 | 1.180 | 0.353 |
| Population Density | 0.017 | 0.004 | <0.001 |
| **Model 4** |  |  |  |
| Tests per 1 Case at 6 Deaths per 1 Million | 0.659 | 0.170 | 0.001 |
| Median Age | 0.005 | 0.346 | 0.988 |
| Country-Level Gross Domestic Product | -0.0002 | 0.0001 | 0.029 |
| Hospital Beds per 1000 | -0.886 | 1.264 | 0.493 |
| Population Density | 0.018 | 0.004 | <0.001 |
| NPI<sup>a</sup> (Sum) | -2.763 | 2.101 | 0.207 |
| **Model 5** |  |  |  |
| Tests per 1 Case at 6 Deaths per 1 Million | 0.659 | 0.170 | 0.001 |
| Median Age | 0.005 | 0.346 | 0.988 |
| Country-Level Gross Domestic Product | -0.0002 | 0.0001 | 0.029 |
| Hospital Beds per 1000 | -0.886 | 1.264 | 0.493 |
| Population Density | 0.018 | 0.004 | <0.001 |
| NPI<sup>b</sup> (Average) | -0.138 | 10.503 | 0.207 |

<sup>a</sup>Ordinary least-squares regression with clustering on a spatial unit (Country)
<sup>b</sup>Measured Country-level Non-Pharmaceutical Interventions (NPI) in place between 6 Deaths per 1 Million and 15 Deaths per 1 Million – NPI included confinement; school/work closures; event cancellations; travel restrictions; and health practices (mask wearing, contact tracing)

Figures 3A-3E indicate heatmap plots corresponding to all effect estimates for Models 1-5, respectively. Shading indicates an association, while varying hues designate the effect size of this association, as noted in the heatmap legend. Across all five models, tests per case at X deaths per million (where X ranges from 6 to 18) is associated with time to Y deaths per million (where Y ranges from 7 to 25), after adjusting for the respective model covariates. In all cases, more testing per case is predictive of a longer time to increased deaths per million.

**Figure 3A-E.**
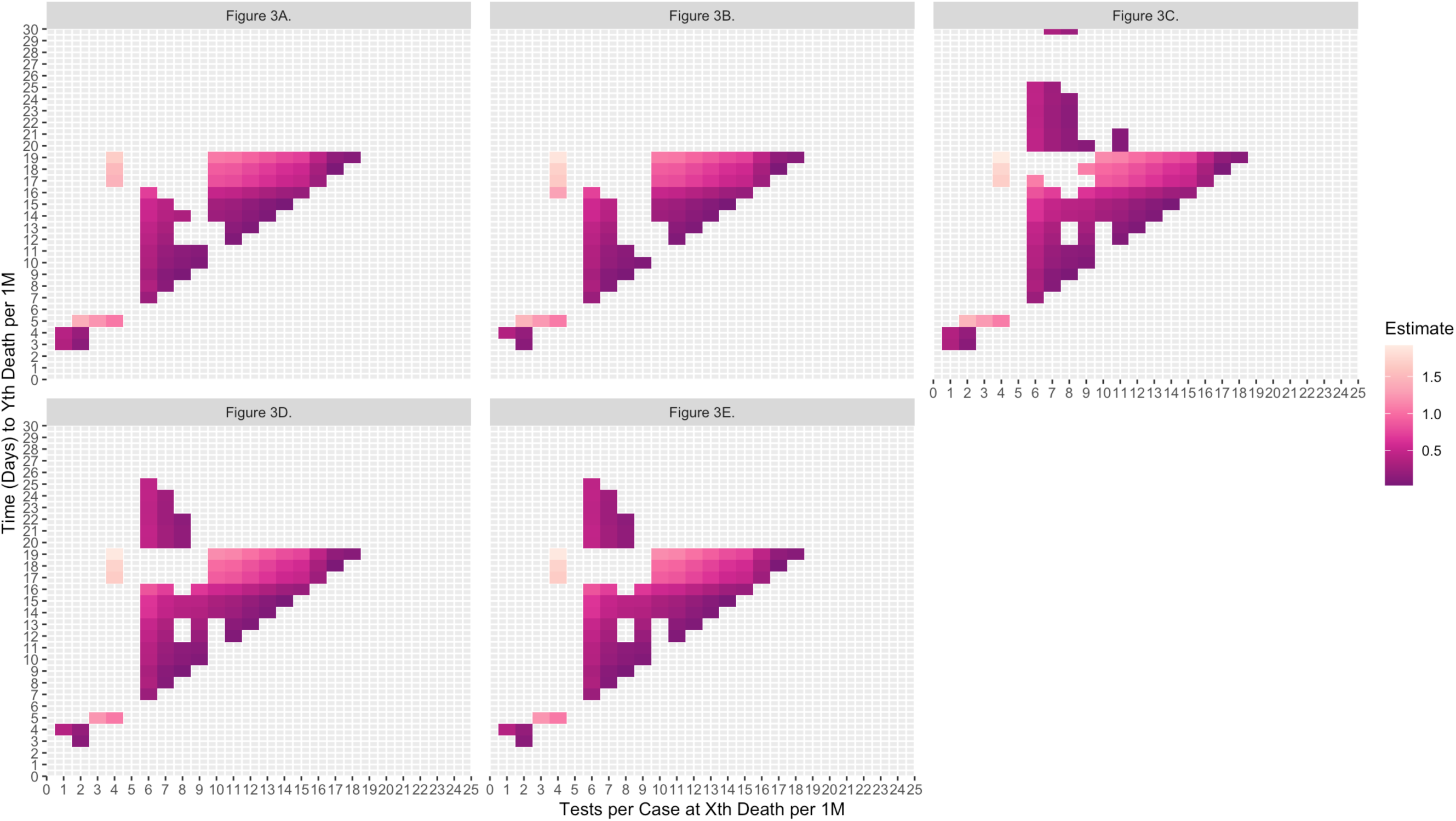
Heatmap Plots of OLS Regression Estimates (For Models of Tests per COVID-19 Case at X^th^ Deaths per Million and Time (Days) to Y^th^ Deaths per Million) Across 27 Countries^a-e^. ^a^Figure 3A. Adjusted for 2 covariates (Median Age, GDP) ^b^Figure 3B. Adjusted for 3 covariates (Median Age, GDP, Hospital Bed Capacity) ^c^Figure 3C. Adjusted for 4 covariates (Median Age, GDP, Hospital Bed Capacity, Population Density) ^d^Figure 3D. Adjusted for 5 covariates (Median Age, GDP, Hospital Bed Capacity, Population Density, NPI_Sum) ^e^Figure 3E. Adjusted for 5 covariates (Median Age, GDP, Hospital Bed Capacity, Population Density, NPI_Average)

## DISCUSSION

We find evidence that early country-level testing capacity is associated with later country-level COVID-19 mortality outcomes, after adjusting for median age, GDP, hospital bed capacity, population density, and NPIs, across 27 countries. More tests per case at an earlier timepoint is associated with increased time to reach later mortality thresholds. Thus, early testing capacity may predict future COVID-19 mortality outcomes. The findings indicate that early, more aggressive administration of diagnostic testing, as realized in certain countries, may have helped slow the accrual of COVID-19 deaths.

There is some existing literature that have addressed the mortality impacts of country-level COVID-19 testing capacity. Leffler et al. (2020) explored the association between viral testing policy and COVID-19 per-capita mortality outcomes across 200 countries, up to May 9, 2020. They operationalized country-level viral testing policy score on the following categorical scale: no implemented policy (0); testing for individuals who are symptomatic with previous exposure (1); testing for individuals who are symptomatic with or without previous exposure (2); or open to all (3). The authors found no association between viral testing policies and COVID-19 mortality outcomes, using linear regression methods.^14^ Similarly, Chaudhry et al. (2020) explored the association between country-level widespread testing, operationalized as tests per million, and COVID-19 per capita mortality across 50 countries with the highest case-load as of April 1, 2020.^12^ They also found no association between country-level wide-spread testing and COVID-19 per capita mortality, using negative binomial regression methods. In contrast, our study, aided in part by an extended study period, finds an association between early country-level testing capacity and later COVID-19 mortality outcomes; however, our study differs in that we measure testing capacity as tests administered per case identified. This measure quantifies the number of tests a country is willing to process to find a single case and is thus an estimate of the testing aggressiveness adopted by a country. Thus, our analysis supplements current research in that it demonstrates that there may be a benefit of more abundant testing relative to case numbers.

Here we use tests per case to estimate the time to particular mortality thresholds. We specifically focus on mortality rates, as tests per case is not an optimal metric for estimating time to cumulative case levels given that more aggressive testing will ascertain a higher fraction of infections as cases. Countries with more active testing strategies may capture a greater number of asymptomatic or paucisymptomatic infections as a result of their more abundant testing and tracing capacity. As such, higher tests per case may slow the accrual of COVID-19 deaths by supporting more isolation and quarantine measures; and thus, better inhibiting further transmission of the virus in the community. A robust country-level testing capacity may also enable earlier diagnosis and treatment of symptomatic cases and improve patient outcomes; thereby, also reducing the number of deaths related to COVID-19.

The major strengths of this study are that we chose to incorporate a time component when measuring mortality outcomes, by operationalizing the outcome as the time interval (in days) to reach a certain country-level mortality threshold. In using a time to threshold measure, we were able to determine whether stronger testing capacity at the onset of the pandemic may have yielded a sustained impact on later mortality outcomes. Furthermore, we adjusted for potential confounders, such as the timing of NPIs, to assess the public health measures that were in place during a particular period. NPIs encompassed important measures such as masking policies, that have been associated with reduced rates of SARS-CoV-2 infection.^15^ We additionally assessed a longer time frame of analysis compared to existing studies, by exploring the association of testing capacity and mortality up until September 30, 2020, utilizing seven months of longitudinal data.

There were certain limitations in our study design approach. We did not identify the type of diagnostic test used in each country. Countries may have had differential access to rapid antigen tests (RAT), or reverse transcription polymerase chain reaction (RT-PCR) tests. These differences are important given that RT-PCR yields higher sensitivity and specificity than RAT, although RAT is relatively quick to administer. We additionally did not account for the type of healthcare system implemented in a country: healthcare access may influence an individual’s access to COVID-19 testing. We also coarsely categorized the NPI variable, which quantified the number of NPIs in place; this variable did not include information regarding the stringency of a specific NPI measure or public compliance. Lastly, the study may have yielded limited generalizability, as findings may only correspond to the countries used in this analysis, during this specific period.

Future directions for research may address using a longer time span to determine if this association between early testing capacity and later mortality outcomes continues to hold at longer time intervals. It may also be helpful to explore whether hospitalization rates per capita play a mediating role between country-level testing capacity and mortality rates per capita in order to assess if higher positivity rates may overrun the capacity of the healthcare system, which may increase COVID-19 mortality rates. Future analyses may also consider the effects of new therapeutics and vaccines in preventing COVID-19 deaths, as this may dilute the association between early testing capacity and later COVID-19 mortality outcomes.

Overall, the study helps operationalize the value in investing in, strengthening, and streamlining COVID-19 testing infrastructures so that populations have more feasible access to testing in an effort to help mitigate future COVID-19 outcomes. During the early stages of the pandemic (spring 2020), epidemiologists did advocate for the roll-out of national testing strategies, including centrally-located testing facilities, increased access for COVID-19 test kits, and the mobilization of community resources, such as specialized advisory groups.^16-17^ These approaches should continue to be examined and prioritized to increase the number of administered tests per case identified on a country-level during the course of the pandemic; this may assist early identification of the SARS-CoV-2 infection, may prevent further spread of the virus, and help mitigate future COVID-19 mortality rates.

## Data Availability

The data that support the findings of this study are publicly available in the 'Our World in Data' database [https://ourworldindata.org/coronavirus], and the 'Worldwide Non-pharmaceutical Interventions Tracker for COVID-19' database [https://ibm.github.io/wntrac/dataset].

https://ourworldindata.org/

https://ibm.github.io/wntrac/dataset/

## TABLES & FIGURES

**Figure Suppl 1A-E.**
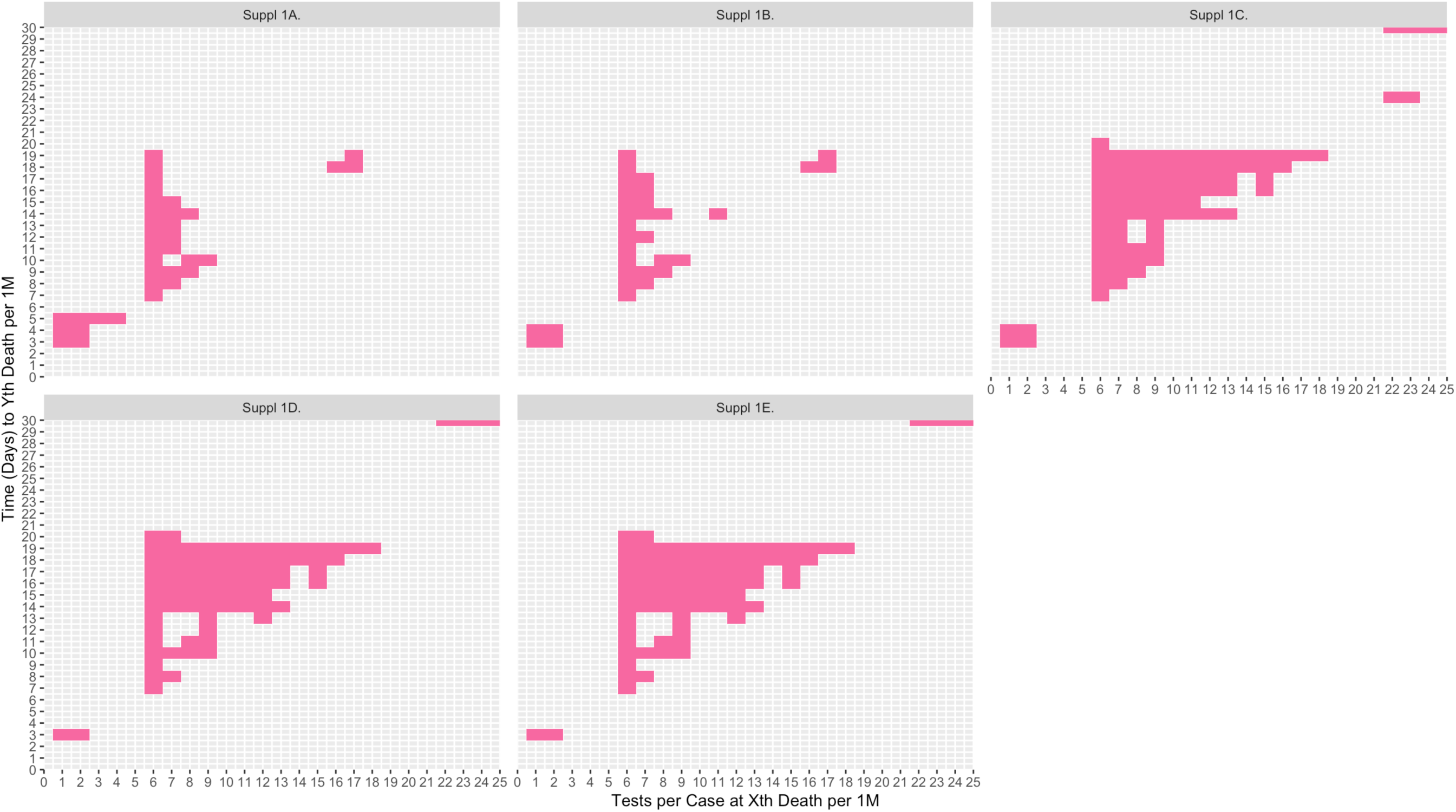
Bootstrap Plots of OLS Regression Estimates (For Models of Tests per COVID-19 Case at X^th^ Deaths per Million and Time (Days) to Y^th^ COVID-19 Deaths per Million) Across 27 Countries^a-e^. ^a^Figure Suppl 1A. Adjusted for 2 covariates (Median Age, GDP) ^b^Figure Suppl 1B. Adjusted for 3 covariates (Median Age, GDP, Hospital Bed Capacity) ^c^Figure Suppl 1C. Adjusted for 4 covariates (Median Age, GDP, Hospital Bed Capacity, Population Density) ^d^Figure Suppl 1D Adjusted for 5 covariates (Median Age, GDP, Hospital Bed Capacity, Population Density, NPI_Sum) ^e^Figure Suppl 1E. Adjusted for 5 covariates (Median Age, GDP, Hospital Bed Capacity, Population Density, NPI_Average)

## ETHICS APPROVAL STATEMENT

This study does not require ethics review given that it uses publicly available and accessible records without contact with the individuals.

## REFERENCES

1. [dataset] Roser M, Ritchie H, Ortiz-Ospina E, and Hasell J (2020) - “Coronavirus Pandemic (COVID-19)”. Published online at OurWorldInData.org. Retrieved from: ‘https://ourworldindata.org/coronavirus‘ [Online Resource]

2. Moghadas SM, Shoukat A, Fitzpatrick MC, et al. Projecting hospital utilization during the COVID-19 outbreaks in the United States. Proc Natl Acad Sci U S A. 2020 Apr 21;117:9122-26.

3. Salari N, Hosseinian-Far A, Jalali R, et al. Prevalence of stress, anxiety, depression among the general population during the COVID-19 pandemic: a systematic review and meta-analysis. Global Health. 2020 Dec;6:57.

4. Ettman CK, Abdalla SM, Cohen GH, Sampson L, Vivier PM, Galea S. Prevalence of depression symptoms in US adults before and during the COVID-19 pandemic. JAMA Netw Open. 2020 Sep 1;3:e2019686.

5. Laborde D, Martin W, Vos R. Poverty and food insecurity could grow dramatically as COVID-19 spreads. International Food Policy Research Institute (IFPRI), Washington, DC. 2020 Apr 16.

6. Health TL. Food insecurity will be the sting in the tail of COVID-19. Lancet Glob Health. 2020 Jun;8:e737.

7. Davies NG, Kucharski AJ, Eggo RM, et al. Effects of non-pharmaceutical interventions on COVID-19 cases, deaths, and demand for hospital services in the UK: a modelling study. Lancet Public Health. 2020 Jul;5:e375–85

8. Jamwal S, Gautam A, Elsworth J, Kumar M, Chawla R, Kumar P. An updated insight into the molecular pathogenesis, secondary complications and potential therapeutics of COVID-19 pandemic. Life Sci. 2020 Sep 17:257;118105.

9. Kandel N, Chungong S, Omaar A, Xing J. Health security capacities in the context of COVID-19 outbreak: an analysis of International Health Regulations annual report data from 182 countries. Lancet. 2020 Mar 28;395:1047–53.

10. Pettit SD, Jerome KR, Rouquié D, et al. ‘All In’: A pragmatic framework for COVID-19 testing and action on a global scale. EMBO Mol Med. 2020 Jun 8;12:e12634.

11. Pan A, Liu L, Wang C, Guo H, et al. Association of public health interventions with the epidemiology of the COVID-19 outbreak in Wuhan, China. JAMA. 2020 May 19;323:1915–23.

12. Chaudhry R, Dranitsaris G, Mubashir T, Bartoszko J, Riazi S. A country level analysis measuring the impact of government actions, country preparedness and socioeconomic factors on COVID-19 mortality and related health outcomes. EClinicalMedicine. 2020 Aug 1;25:100464.

13. [dataset] Suryanarayanan P, Tsou CH, Poddar A, et al. WNTRAC: Artificial Intelligence Assisted Tracking of Non-pharmaceutical Interventions Implemented Worldwide for COVID-19. arXiv preprint arXiv:2009.07057. 2020 Sep 2.

14. Leffler CT, Ing EB, Lykins JD, Hogan MC, McKeown CA, Grzybowski A. Association of country-wide coronavirus mortality with demographics, testing, lockdowns, and public wearing of masks. Am J Trop Med Hyg. 2020 Dec;103:2400–11.

15. Wang X, Ferro EG, Zhou G, Hashimoto D, Bhatt DL. Association between universal masking in a health care system and SARS-CoV-2 positivity among health care workers. JAMA. 2020 Jul 14;324:703–4.

16. Peto J. Covid-19 mass testing facilities could end the epidemic rapidly. BMJ. 2020 Mar 22;368:m1163.

17. Peto J, Alwan NA, Godfrey KM, et al. Universal weekly testing as the UK COVID-19 lockdown exit strategy. Lancet. 2020 May 2;395:1420–1.

